# On some general characteristics of the spread of the COVID epidemic

**DOI:** 10.1101/2023.09.22.23295959

**Authors:** Denis Below, Felix G. Mairanowski

## Abstract

An analytical model for calculating the spread of the COVID-19 epidemic is described, and it is shown that the main patterns of epidemic development are determined by three dimensionless complexes representing the ratios of transmission rates, contact limitation due to lockdown, and population vaccination. A comparison of statistical data on the growth of the epidemic in Germany, Berlin and its various neighbourhoods shows that the development of the epidemic depends to a large extent on the ethnic composition of the population. In the same way as it is accepted in some sections of physics, it is proposed to use the methods of similarity theory to investigate the regularities of the emergence and development of the epidemic.

---

The accumulation of many years of continuous statistical data on the spread of COVID-19 in different countries allows us to significantly expand our understanding of various aspects of the emergence and course of the epidemic. More than a hundred different computational models have been proposed to describe the development of the epidemic. Some of them are based on the solution of a system of differential equations of various levels of detail, others are purely stochastic numerical models or analyse statistical data on the number and duration of contacts leading to virus transmission. All these models require the introduction of a number of coefficients for their agreement with observational data, and the number of coefficients, as a rule, increases with the complexity of the proposed models.

We propose a relatively simple model that allows us to obtain analytical solutions. Let us write the initial differential equations of the epidemic model, taking into account the impact of lockdown and mass vaccination on the epidemic spread, as [1]:

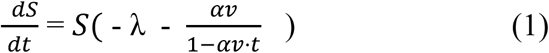

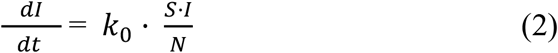

where:

I - the number of infected persons at a given time,

*k*_0_ - coronavirus infection rate (1/day)

N - total population of the area under consideration,

S - the number of susceptible part of the population potentially capable of becoming infected due to contact with infected individuals,

λ - intensity factor of decrease in contacts of infected patients with persons who potentially can get infected by means of quarantine and other preventive measures,

v - population vaccination rate (1/day),

α - is the coefficient of vaccine effectiveness.

In the tradition of mathematical modelling in epidemiology, this model will hereafter be referred to as ASILV for short. This name underlines the main features of this model, namely:

A is an analytical model by which an analytical solution of the system of equations can be obtained, S is the part of the population that is not yet infected, but which could become infected through contact with infected individuals, I is the part of the population that has already been infected at time t, L is the part of the population that is protected from infection by lockdown measures at the time in question, but which could potentially also become infected later if the lockdown conditions change, V is the part of the population that is protected from infection by vaccination, the effectiveness of which α may vary.

Equation (1), defines the change in the number of persons potentially susceptible to the virus under conditions of lockdown and mass vaccination of the population. The denominator in the last summand of equation (1) takes into account that as the proportion of the vaccinated population αv*t increases, the degree of impact of vaccination on the declining epidemic increases. The coefficient of effectiveness α depends on both the type of vaccine and the number of vaccination dose (first or second). We will assume that the maximum vaccination rate will not exceed (αv*t) _max_ ≤ 0.8, i.e. that with an 80% vaccination rate the epidemic cannot develop. This is a natural limitation of the proposed model. However, we have to take into account that some part of the population has already had the disease either explicitly or asymptomatically by the time mass vaccination begins.

The solution to equation (1) is as follows:

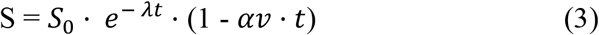

After substituting (3) into (2), solving the resulting equation, transformations and moving to a relative number of infections, we obtain the basic calculation equations.

For the period from outbreak to mass vaccination *t* _*v*_ that is

for t ≤ *t* _*v*_, when *αv =* 0, the solution of equation (2) has the form

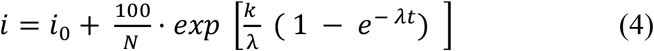

*i* - is the relative number of infected persons per one inhabitant of the settlement in question, as a percentage,

*i*_0_ - is the value of i at the initial moment of the calculation period,

k - is the transmission rate coefficient for the settlement with a population of N, which is calculated by the formula :

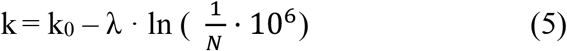

The k_0_ coefficient also depends on the transmissibility of the virus strain responsible for the epidemic spread during the period under consideration.

In contrast to previously developed models of this type, in our model there is no inverse relationship between the values of I and S, i.e. under lockdown conditions and provided that I ≪ S (as a rule I ≤ 0.2 S), we can assume that the number of potentially susceptible individuals does not vary with the number of infections and depends only on the severity of the lockdown It would seem that the condition that I ≪ S under conditions of mass vaccination sharply limits the application of this model. But, as it was found, the effect of vaccination decreases sharply with time. In addition, the emergence of new strains allows the virus to overcome the protective barriers of vaccination. Therefore, it can be considered that this computational model is also applicable in conditions of mass vaccination of the population.

As a result of solving the system of equations and transfer to the relative number of infections, we obtain the basic calculation relation for t ≥ *t* _*v*_ :

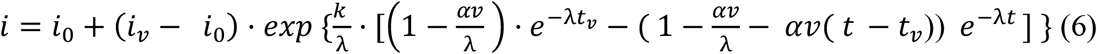

Where *i = i*_*v*_ at t *= t* _*v*_

*i* - is the relative number of infected persons per one inhabitant of the settlement in question, as a percentage,

*i*_0_ - is the value of i at the initial moment of the calculation period.

The process of epidemic development, according to the developed model, is defined by three dimensionless parameters: k /λ, αv/λ and λt. All variables that make up these complexes have dimensionality 1/s and characterise the intensities of individual processes: k - infection, λ - reduction of infections as a result of lockdown and αv - vaccination. Each of these processes depends on many factors, some of them can be regulated, and others cannot. The parameter k, which depends on virus strain, population size and climatic conditions, is not controllable. The other two parameters depend on the effectiveness of lockdown and vaccination respectively, i.e. they can and should be administratively regulated.

At the initial stage of the epidemic, when effective vaccination methods have not yet been developed, the calculations are based on the ratio (4). A comparison of the results of calculations for the first and second waves of the epidemic, carried out according to (4) for Berlin and its different districts, as well as for Germany, showed a good agreement with the statistical data. By calculating the coefficient k according to formula (5) and changing the coefficient λ, the value of which should depend on the level of severity of the lockdown, the agreement of the corresponding curves was obtained, with a correlation value between them of 0.93 and 0.98. However, an important general pattern was simultaneously established: some curves of relative growth of the epidemic practically coincide.

Figure 1 shows the results of the statistics on the relative growth of infections during the second wave of the epidemic.

**Fig. 1.**
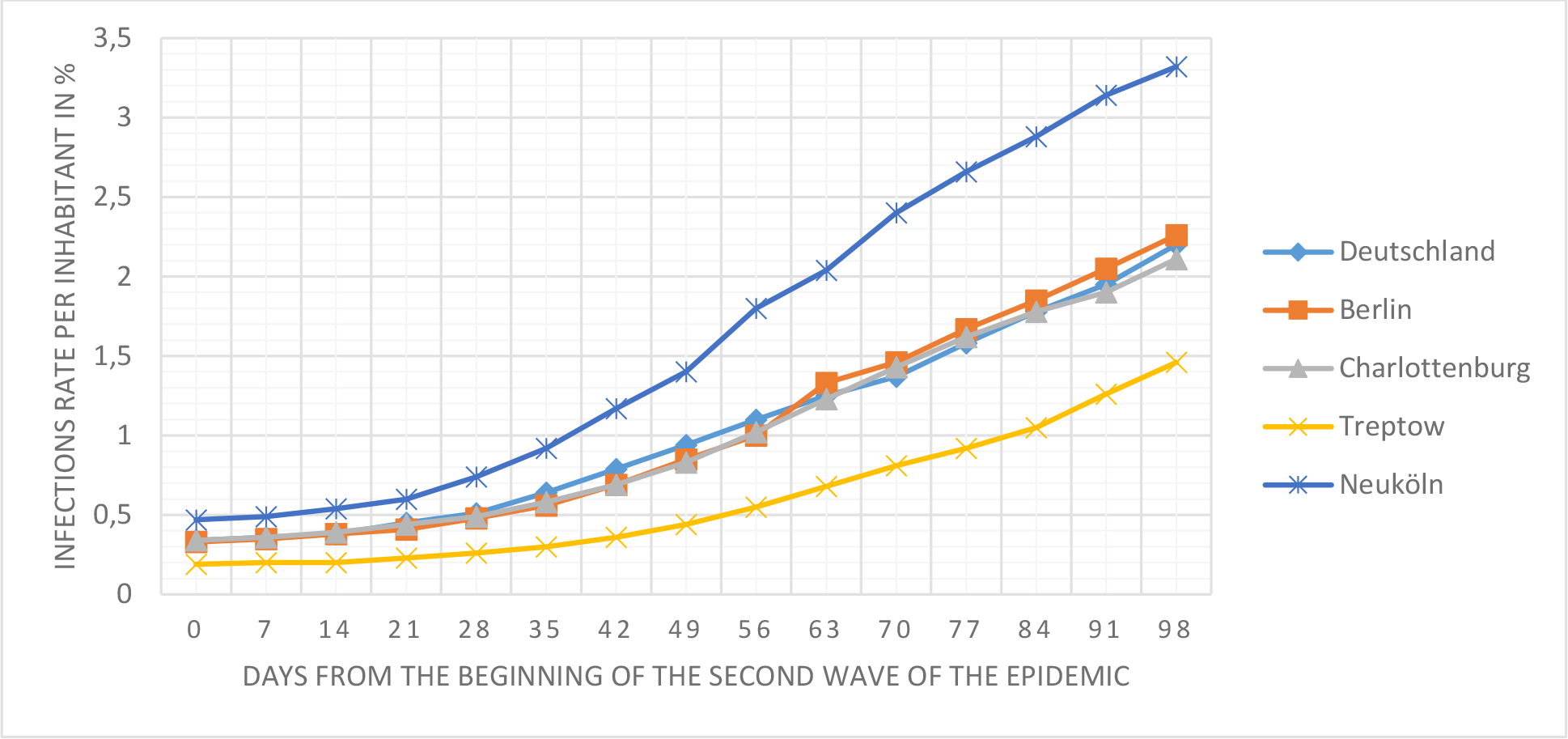
The depends of infection rate on foreigners from countries OIC

As can be seen from this figure, there is a great unevenness in the number of infected persons across the city. Thus, while in some neighbourhoods, such as Treptow-Köpenik, the maximum increase in the number of infected persons does not exceed 50 persons per day, in the Neuköln neighbourhood it reaches 300 persons per day and more. Such high numbers in this area can be explained by certain behavioural characteristics of the inhabitants of this area, numerous migrants from the member countries of the Organisation of the Islamic Conference (OIC) (Turkey, Arab States, some African countries). Thus, outbreaks of the viral disease were often recorded after crowded celebrations of various events, which are traditional for the inhabitants of these countries. At the same time, it was impossible to identify the chain of participants of these celebrations, which did not allow preventing further mass infection.

It is important to emphasise that the growth curves of not only the first but also the second wave of the epidemic for the district of Charlottenburg practically coincide with the curves for Berlin and for Germany as a whole. The relative numbers of OIC residents for Charlottenburg, Berlin and Germany are 12 per cent, 11 per cent and about 10 per cent respectively. For Neuköln with the maximum number of infected people this figure exceeds 20%, for Treptow-Köpenik with the minimum number of diseases it is about 4%. 20%, for Treptow-Köpenik with the minimum number of diseases it is about 4%. The application of the proposed model allowed us to determine the dependence of the coefficient λ on the relative number of residents with emigrant roots - natives of OIC countries [2]: For Neuköln with the maximum number of infected people this figure exceeds 20%, for Treptow-Köpenik with the minimum number of diseases it is about 4%. 20%, for Treptow-Köpenik with the minimum number of diseases it is about 4%. The application of the proposed model allowed us to determine the dependence of the coefficient λ on the relative number of residents with emigrant roots - natives of OIC countries [2]:

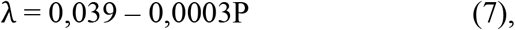

where P is the relative number of citizens - natives of OIC countries. The correlation coefficient between the statistical data and the ratio (7) is 0.89%.

Apparently, foreigners from member countries of Islamic countries have closer family and friendship ties, which can contribute to more intensive transmission of the virus. It would be more correct to relate the intensity of transmission to certain psychological characteristics according to the theory on the behavioural immune system (BIS) .However, the study of these problems is at the initial stage and special comprehensive psychological studies are needed to solve them. In order to study the influence of the main factors determining the rate of epidemic growth, hence having the maximum effect on the value of the λ coefficient, it is necessary to perform additional studies on the peculiarities of infra-texture and socio-demographic characteristics for each of Berlin’s districts. At the same time, it is necessary to take into account the large variability of these factors within most districts, in which regions with a homogeneous structure can be additionally distinguished

Interestingly, ethnic differences have had a significant impact on the epidemic in New York City, where the rate of COVID infection among Latinos is more than twice as high as among whites. Apparently, these differences are also related to the behavioural characteristics of different ethnic groups of the population.

In order to exclude the influence of the previous wave, the difference between the current value of the number of infected people and their values at the time of the second wave is used. Using this technique, observations of the second wave of the epidemic in New York, Germany and Berlin were compared. These results are presented in Fig. 2. As can be seen from this figure, all three curves practically coincide up to a certain point in time. Only about 90 days after the beginning of this epidemic wave, the growth of the infection in New York became significantly higher than in Berlin and Germany. That means that the passage of the third wave of the epidemic for New York differs significantly from that of Berlin.. Apparently, the increase in the epidemic during this period of time could be explained by the increase in human contact between Christmas and New Year’s Eve. During this time period, there were also deviations of the calculated data from the fixed data for Germany and Berlin. However, the magnitude of these deviations is significantly lower than for New York. Thus, it can be assumed that the intensity of contacts in New York is higher than in Germany and Berlin.

**Fig. 2.**
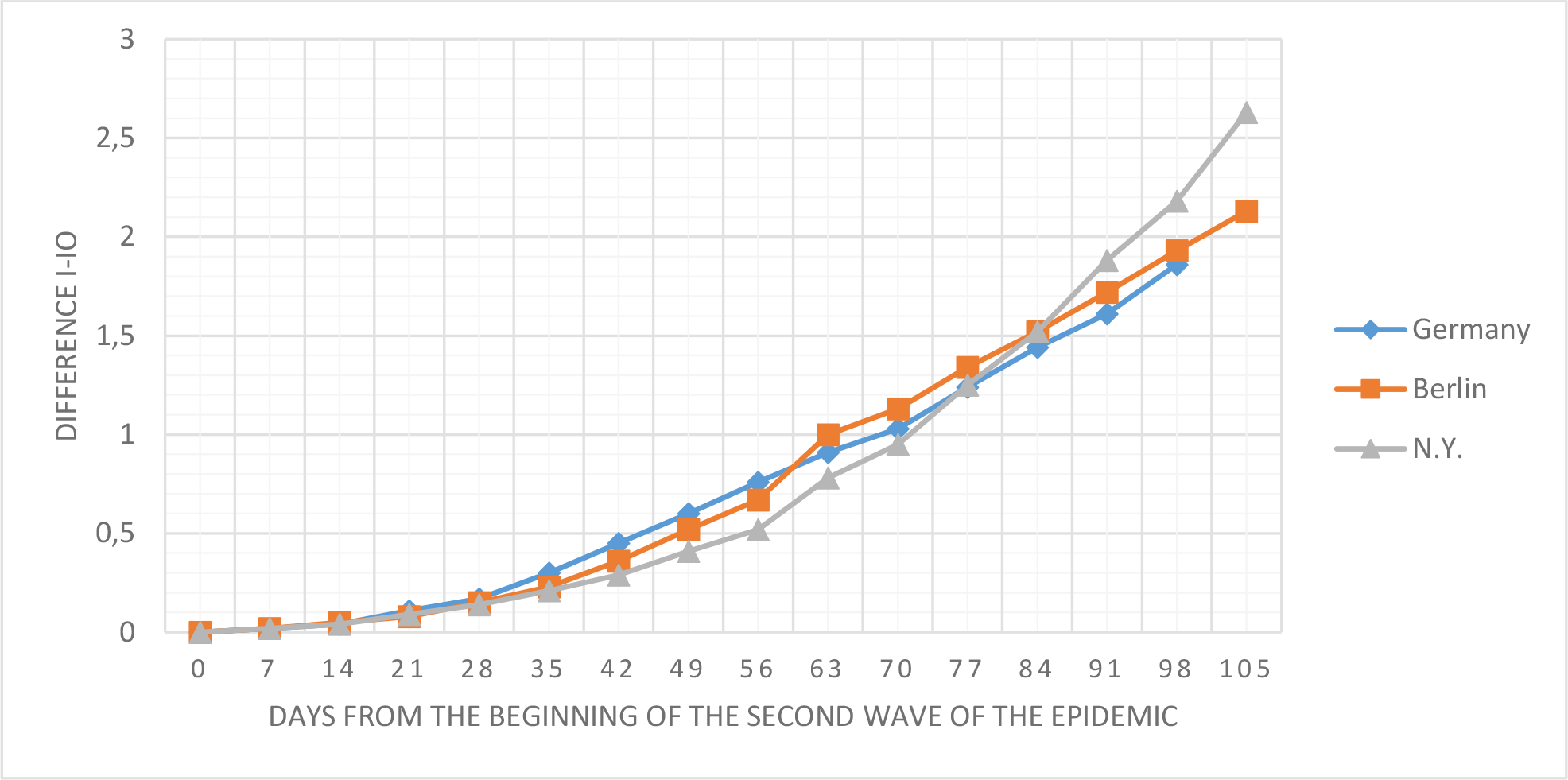
Changes in the number of infections compared to their numbers at the beginning of the second wave

If we consider that the strongest influence on the development of the epidemic and, consequently, on the value of the coefficient λ in our calculation model is exerted by the intensity of personal contacts, it is natural to distinguish population groups according to their age. Fig. 3 shows the results of our research on the change in the coefficient λ depending on age [3]

**Fig. 3.**
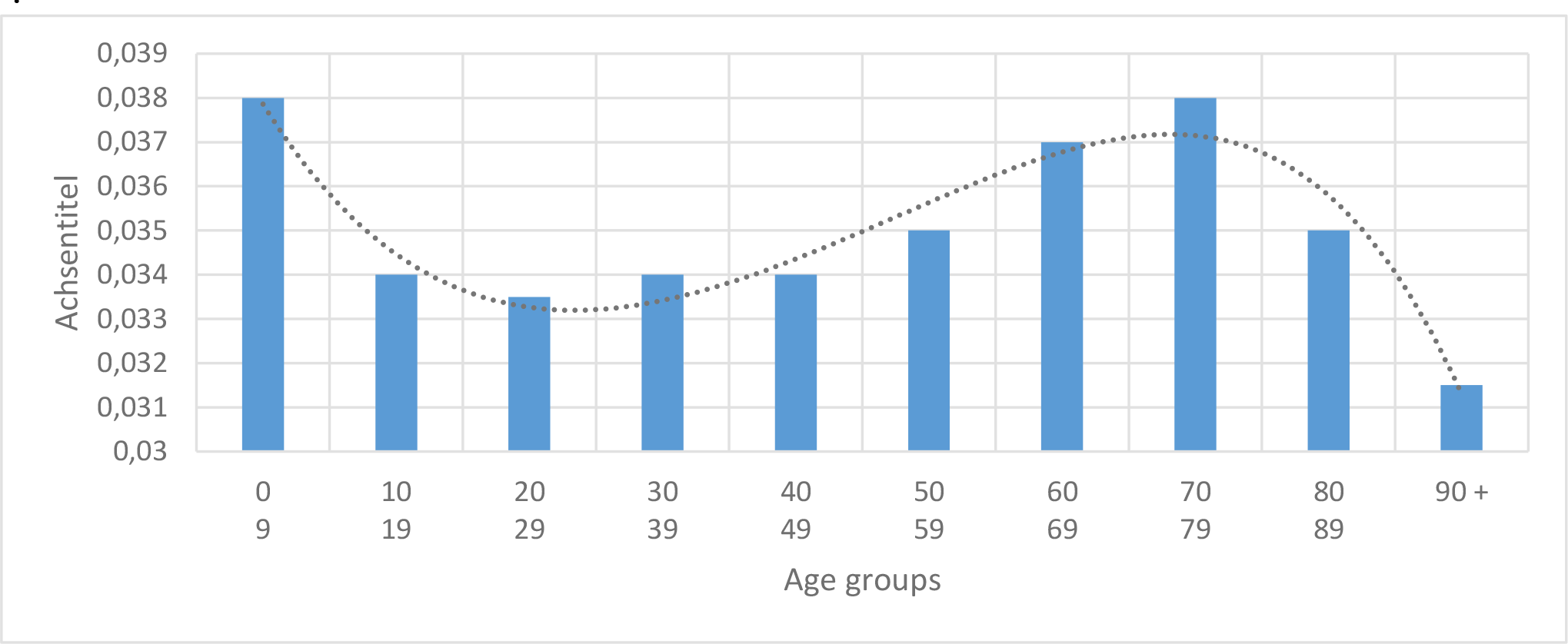
Dependence of the coefficient on the age of different population groups in Berlin

In particular, one of these factors may be differences in the age distribution of the population in different districts. As was already established in our previous work, the maximum percentage of infected people are young people between the ages of 20 and 29 who are the most socially active. The disease rate in this age group reaches 1.5%. School-age children and people in the 30 to 50 age group are slightly weaker, slightly more than 1%. This conclusion was made based on the study of the epidemic in its initial phase. At this stage of the epidemic the differences between the relative numbers of infected persons from 10 to 60 years old have slightly decreased. A more detailed analysis shows that more than 50% of all infected people (over 41,000) belong to the age group of 20 to 50 years old. The maximum percentage of those infected (3.36%) belongs to the most active age group of 20 to 24 years old. As for the elderly people 90+, although the relative number of those infected is quite high, the absolute number of people of this age group for Berlin is just over 1,000 people. In this regard, some doubts can be raised about the advisability of first vaccinating the elderly. Given that to date there is no reliable information about the possible effects of vaccination especially on immunocompromised individuals, it may be more appropriate to vaccinate the 18-30 age group in order to minimize the speed of the epidemic. Naturally, this most active age group generally has stronger immunity, which reduces the possibility of negative effects associated with vaccination.

Different age groups are characterised not only by different attitudes towards meeting the lockdown conditions, but also by different responses to vaccination requirements. In Berlin, for example, the vaccination rate for the age group 20-29 years was about 0.002 1/day, whereas for populations 30 and above it is about 3 times higher, and for the elderly over 90 years of age it reaches a value of almost 0.01 1/day. Thus, in general, the trends of λ and v parameters coincide.

Using indicative data, an attempt was made to relate this parameter to the level of attenuation of transmission intensity due to lockdown. A coefficient L is introduced to estimate the degree of attenuation of transmission intensity. In the absence of a lockdown, the value of L is maximum, i.e. equal to 1. When the value of L decreases to minimum values equal to 0.04 (i.e. when the intensity of virus transmission decreases by 25 times), it is assumed that the epidemic is completely suppressed. Under these conditions, an approximating formula was obtained [4]:

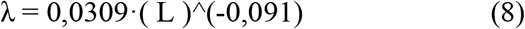

Given the uncertainty of the data on the stringency of the transmission restriction requirements, the λ coefficient was selected at this stage in the epidemic spread calculations by comparing calculated and statistical data.

Control calculations using only a single empirical coefficient showed high accuracy. The calculated curves for such different regions as Germany, Berlin and its districts, Scotland, New York, Israel, South Africa, etc. agree well with the corresponding statistical data. Moreover, good agreement between the fixed and calculated data is also found for individual age groups of the population.

In addition to the proposed model, it is of great importance that regions with completely different areas and populations have a uniform pattern of epidemic development. This is evidenced, in particular, by the data in Figure 1, according to which the epidemic development curves for Germany, Berlin and the relatively small region of Charlottenburg coincide completely. An analogy with the theory of similarity, which is widely used in practical research in various sections of physics (see, e.g., [5].). In the same way as, for example, small-scale modelling methods are widely used in solving many practical problems related to heat and mass transfer and hydromechanics, it is also possible to carry out studies of epidemic reduction within the limits of some relatively small areas, being sure that the results obtained can be transferred to wider areas. Thus, for example, within a relatively small area there is a real possibility to perform a detailed analysis of the level of limitation of the number and duration of personal contacts, compliance with the mask regime, hygiene standards, etc.

Further development of similarity methods in epidemiology opens up the possibility of applying new effective methods of studying the patterns of infection spread and will allow to accelerate the development of recommendations for optimising the fight against epidemics.

## Data Availability

All data produced in the present study are available upon reasonable request to the authors.

## Notes

### Competing Interest Statement

The authors have declared no competing interest.

### Funding Statement

This study did not receive any funding.

